# Associations of DMT therapies with COVID-19 severity in multiple sclerosis

**DOI:** 10.1101/2021.02.08.21251316

**Authors:** Steve Simpson-Yap, Edward De Brouwer, Tomas Kalincik, Nick Rijke, Jan Hillert, Clare Walton, Gilles Edan, Yves Moreau, Tim Spelman, Lotte Geys, Tina Parciak, Clément Gautrais, Nikola Lazovski, Ashkan Pirmani, Amin Ardeshirdavani, Lars Forsberg, Anna Glaser, Robert McBurney, Hollie Schmidt, Arnfin Bergmann, Stefan Braune, Alexander Stahmann, Rodden Middleton, Amber Salter, Robert J. Fox, Anneke van der Walt, Helmut Butzkueven, Raed Al-Roughani, Serkan Ozakbas, Juan I Rojas, Ingrid van der Mei, Nupur Nag, Rumen Ivanov, Guilherme Sciascia do Olival, Alice Estavo Dias, Melinda Magyari, Doralina Guimarães Brum, Maria Fernanda Mendes, Ricardo Alonso, Richard Nicholas, Johana Bauer, Anibal Chertcoff, Ana Zabalza, Georgina Arrambide, Alexander Fidao, Giancarlo Comi, Liesbet M. Peeters

**Affiliations:** CORe, Department of Medicine, The University of Melbourne, Australia; Neuroepidemiology Unit, Melbourne School of Population & Global Health, The University of Melbourne, Australia; Menzies Institute for Medical Research, University of Tasmania, Australia; ESAT-STADIUS, KU Leuven, Belgium; Melbourne MS Centre, Department of Neurology, Royal Melbourne Hospital, Australia; MS International Federation, United Kingdom; Department of Clinical Neuroscience, Swedish MS Registry, Sweden; Department of Neurology, CHU Pontchaillou, France; Karolinska Institutet, Sweden; Biomedical Research Institute - Data Science Institute, Hasselt University, Belgium; Department of Medical Informatics, University Medical Center Göttingen, Germany; Department of Computer Science and AI, KU Leuven, Belgium; QMENTA, Spain; iConquerMS People-Powered Research Network, Accelerated Cure Project for MS, United States of America; NeuroTransData Study Group, NeuroTransData, Germany; German MS-Register by the National MS Society, MS Forschungs- und Projektentwicklungs-gGmbH, Germany; UK MS Register, Swansea University, United Kingdom; COViMS, United States of America; Division of Biostatistics, Washington University in St Louis, United States of America; Mellen Center for Multiple Sclerosis, Cleveland Clinic, United States of America; Department of Neuroscience, Central Clinical School, Monash University, Australia; Dokuz Eylul University, Turkey; Division of Neurology, Department of Medicine, Amiri Hospital, Kuwait; Neurology Department, Hospital Universitario de CEMIC, Argentina; RELACOEM, Argentina; The Australian MS Longitudinal Study, Menzies Institute for Medical Research, University of Tasmania, Australia; Bulgarian SmartMS COVID-19 Dataset, Bulgaria; ABEM - Brazilian MS Patients Association, Brazil; The Danish Multiple Sclerosis Registry, Departement of Neurology, University Hospital Rigshospitalet, Denmark; Universidade Estadual Paulista, Unesp, Faculdade de Medicina, Botucatu, Brazil; REDONE.br – Brazilian Registry of Multiple Sclerosis and Neuromyelitis Optica Spectrum Disorders, Brazil; Irmandade da Santa Casa de Misericórdia de São Paulo, Brazil; Multiple Sclerosis University Center, Ramos Mejia Hospital – EMA, Argentina; Imperial College London, United Kingdom; Swansea University, United Kingdom; Mental Health Area, EMA, Argentina; MS and Demyelinating Diseases. Hospital Británico de Buenos Aires, EMA, Argentina; Servei de Neurologia-Neuroimmunologia. Centre d’Esclerosi Múltiple de Catalunya, (Cemcat). Vall d’Hebron Institut de Recerca, Vall d’Hebron Hospital Universitari. Universitat Autònoma de Barcelona, Spain; Institute of Experimental Neurology, Ospedale San Raffaele, Italy

**Author notes:** Corresponding author: Professor Dr Liesbet M Peeters, Biomedical Research Institute & Data Science Institute, Hasselt University, Belgium.

## Abstract

**Background:** People with multiple sclerosis (MS) are a vulnerable group for severe COVID-19, particularly those taking immunosuppressive disease-modifying therapies (DMTs). We examined the characteristics of COVID-19 severity in an international sample of people with MS.

**Methods:** Data from 12 data-sources in 28 countries were aggregated. Demographic and clinical covariates were queried, alongside COVID-19 clinical severity outcomes, hospitalisation, admission to ICU, requiring artificial ventilation, and death. Characteristics of outcomes were assessed in patients with suspected/confirmed COVID-19 using multilevel mixed-effects logistic regression.

**Results:** 657 (28.1%) with suspected and 1,683 (61.9%) with confirmed COVID-19 were analysed. Older age, progressive MS-phenotype, and higher disability were associated with worse COVID-19 outcomes. Compared to dimethyl fumarate, ocrelizumab and rituximab were associated with hospitalisation (aOR=1.56,95%CI=1.01-2.41; aOR=2.43,95%CI=1.48-4.02) and ICU admission (aOR=2.30,95%CI=0.98-5.39; aOR=3.93,95%CI=1.56-9.89), though only rituximab was associated with higher risk of artificial ventilation (aOR=4.00,95%CI=1.54-10.39). Compared to pooled other DMTs, ocrelizumab and rituximab were associated with hospitalisation (aOR=1.75,95%CI=1.29-2.38; aOR=2.76,95%CI=1.87-4.07) and ICU admission (aOR=2.55,95%CI=1.49-4.36; aOR=4.32,95%CI=2.27-8.23) but only rituximab with artificial ventilation (aOR=6.15,95%CI=3.09-12.27). Compared to natalizumab, ocrelizumab and rituximab were associated with hospitalisation (aOR=1.86,95%CI=1.13-3.07; aOR=2.88,95%CI=1.68-4.92) and ICU admission (aOR=2.13,95%CI=0.85-5.35; aOR=3.23,95%CI=1.17-8.91), but only rituximab with ventilation (aOR=5.52,95%CI=1.71-17.84). Importantly, associations persisted on restriction to confirmed COVID-19 cases. No associations were observed between DMTs and death.

**Conclusions:** Using the largest cohort of people with MS and COVID-19 available, we demonstrated consistent associations of rituximab with increased risk of hospitalisation, ICU admission, and requiring artificial ventilation, and ocrelizumab with hospitalisation and ICU admission, suggesting their use may be a risk factor for more severe COVID-19.

## Introduction

Disease-modifying therapies (DMTs) that act by immunomodulatory/immunosuppressive mechanisms are a mainstay of treatment of multiple sclerosis (MS) but can increase infection susceptibility^1^. Of current concern is the novel coronavirus, SARS-CoV-2, the cause of Coronavirus Disease of 2019 (COVID-19)^2^. It remains unclear whether people with MS, especially those treated with immunosuppressive DMTs, are more susceptible to more severe COVID-19. Case-series^3-6^ and cohort^7-9^ studies suggest that MS in general does not increase risk for developing severe COVID-19, but comorbidities, age, sex, progressive MS-phenotype, and higher disability do^7, 9^.

Some studies have also identified associations of certain DMT classes, particularly B-cell-depleting DMTs, with COVID-19 severity. However, this varied among studies, potentially due to limited sample size and regional differences in DMT usage^7, 9^. Large and geographically inclusive cohorts are required to assess the risk of severe COVID-19 for specific MS DMTs. Accordingly, we established a global data-sharing initiative^10^ to investigate characteristics of COVID-19 severity in people with MS.

## Methods

### Data-sources

This study was approved by the ethical committee of Hasselt University [CME2020/025]. Individual data-sources obtained additional ethics approval as required.

Data from a core questionnaire regarding COVID-19, and relevant patient demographic and neurological information, were reported by treating clinicians, as described previously^10^. Data were entered in three fashions: 1) “Direct entry to central platform”; 2) “Patient-level data sharing via participating registries/cohorts”, where MS registries and cohorts are regularly invited to share and upload their COVID-19 core dataset into the central data platform; and 3) “Aggregated data sharing via participating registries/cohorts”, where some registries do not share patient-level data, but share aggregated results from specific queries. Multidimensional contingency tables from 12 different data-sources were merged and then a combined anonymised dataset was reconstructed.

Data was entered for a given participant once, but information for that participant could be reentered and this then replacing the original record. This thus made for serial iterations of the analysis dataset, which were analysed over time as the dataset expanded, thus allowing for assessment of temporal consistency of observed associations. Previous iterations of the dataset have been presented^11^ but this article describes only the present iteration of the dataset.

### Definition of variables

COVID-19 status was defined as confirmed, based on a positive diagnostic test, or suspected, based on clinician judgement.

Patient age at the time of reporting was categorised into four groups: 0-17, 18-49, 50-69, and ≥70 years; the 0-17 group was excluded (n=5). MS-phenotype was grouped into relapsing-remitting MS (RRMS) and progressive MS (SPMS, PPMS). Disability was assessed by the Expanded Disability Status Scale (EDSS)^12^ or Neurostatus^13^. Disability was dichotomised into 0-6.0 and >6.0. Comorbidities were queried, including cardiovascular disease, hypertension, diabetes, chronic liver disease, kidney disease, other neurological/neuromuscular disorder, lung disease, or malignant neoplasia. BMI was categorised as non-obese (BMI≤30) and obese (BMI>30). Current smoker status was queried as yes or no. Current DMT use was queried, including alemtuzumab, cladribine, dimethyl fumarate, fingolimod, glatiramer-acetate, beta interferons, natalizumab, ocrelizumab, rituximab, siponimod, teriflunomide, or Other DMT. Due to patient numbers <20 among the suspected/confirmed COVID-19 cases, siponimod (n=12) was aggregated with Other DMT.

### Statistical analysis

Associations with hospitalisation, ICU admission, ventilation, and death were assessed using multilevel mixed-effects logistic regression, random effects grouped by data source, as univariable and adjusted for age, sex, MS-phenotype, and disability. In addition, sensitivity-analyses were completed serially excluding each data-source to assess whether influential data sources underlay associations. Subgroup analyses were also undertaken where data on comorbidities, BMI, and smoking were available, allowing additional adjustment for these covariates. All analyses were complete-case.

For DMTs, individual DMTs were first compared with dimethyl fumarate. Despite leading to lymphopenia in some patients, dimethyl fumarate has not been associated with increased risk of infection^14^, and its biological mechanism of action is unlikely to interfere with the immunological response to SARS-CoV-2^15^, while being common in the sample. Next ocrelizumab and rituximab and the untreated were compared with pooled other DMTs. Finally, ocrelizumab and rituximab were evaluated vs natalizumab to assess ascertainment bias, since natalizumab-treated patients present for infusions every 28-42 days, compared to biannual infusions for anti-CD20 DMTs. Data analyses were carried out using STATA/SE 16.0 (StataCorp, College Station, USA).

## Results

The cohort comprised 2,340 patients, of whom 657 (28.1%) had suspected COVID-19 and 1,683 (71.9%) had confirmed COVID-19. Among suspected/confirmed COVID-19 cases, which was the primary analysis dataset, 20.9% were hospitalised, 5.4% admitted to ICU, 4.1% required artificial ventilation, and 3.2% died. Proportions were slightly higher among confirmed COVID-19 cases (Table 1).

**Table 1.** Cohort characteristics.

|  | Suspected & confirmed (n=2340) | Confirmed (n=1683) |
| --- | --- | --- |
| **COVID-19 status** |  |  |
| Suspected | 657 (28.1%) | 0 (0.0%) |
| Confirmed | 1683 (71.9%) | 1683 (100.0%) |
| **Hospitalisation** |  |  |
| No | 1705 (72.9%) | 1144 (68.0%) |
| Yes | 489 (20.9%) | 453 (26.9%) |
| Missing | 146 (6.2%) | 86 (5.1%) |
| **ICU admission** |  |  |
| No | 1839 (78.6%) | 1355 (80.5%) |
| Yes | 127 (5.4%) | 122 (7.2%) |
| Missing | 338 (14.4%) | 199 (11.8%) |
| **Ventilation** |  |  |
| No | 1755 (75.0%) | 1316 (78.2%) |
| Yes | 97 (4.1%) | 91 (5.4%) |
| Missing | 452 (19.3%) | 269 (16.0%) |
| **Death** |  |  |
| No | 2053 (89.1%) | 1491 (88.6%) |
| Yes | 73 (3.2%) | 65 (3.9%) |
| Missing | 178 (7.7%) | 120 (7.1%) |
| **Sex** |  |  |
| Male | 637 (27.2%) | 469 (27.9%) |
| Female | 1702 (72.7%) | 1213 (72.1%) |
| Missing | 1 (0.0%) | 1 (0.1%) |
| **Age** |  |  |
| 18-<50 | 1505 (64.3%) | 1032 (61.3%) |
| 50-<70 | 740 (31.6%) | 571 (33.9%) |
| 70+ | 83 (3.5%) | 72 (4.3%) |
| Missing | 12 (0.5%) | 8 (0.5%) |
| **EDSS** |  |  |
| 0-6 | 1837 (78.5%) | 1275 (75.8%) |
| >6 | 438 (18.7%) | 368 (21.9%) |
| Missing | 65 (2.8%) | 40 (2.4%) |
| **Has comorbidities** |  |  |
| No | 984 (43.5%) | 745 (45.5%) |
| Yes | 920 (40.7%) | 707 (43.2%) |
| Missing | 357 (15.8%) | 184 (11.3%) |
| **BMI** |  |  |
| No | 820 (36.3%) | 656 (40.1%) |
| Yes | 437 (19.3%) | 376 (23.0%) |
| Missing | 1004 (44.4%) | 604 (36.9%) |
| **Current smoker** |  |  |
| No | 1627 (72.0%) | 1266 (77.4%) |
| Yes | 166 (7.3%) | 115 (7.0%) |
| Missing | 468 (20.7%) | 255 (15.6%) |
| **DMT** |  |  |
| Untreated | 284 (12.1%) | 231 (13.7%) |
| Alemtuzumab | 31 (1.3%) | 26 (1.5%) |
| Cladribine | 29 (1.2%) | 17 (1.0%) |
| Dimethyl fumarate | 275 (11.8%) | 199 (11.8%) |
| Fingolimod | 200 (8.5%) | 143 (8.5%) |
| Glatiramer acetate | 87 (3.7%) | 70 (4.2%) |
| Interferon | 124 (5.3%) | 85 (5.1%) |
| Natalizumab | 221 (9.4%) | 164 (9.7%) |
| Ocrelizumab | 471 (20.1%) | 365 (21.7%) |
| Rituximab | 258 (11.0%) | 142 (8.4%) |
| Teriflunomide | 97 (4.1%) | 75 (4.5%) |
| Other DMT | 70 (3.0%) | 47 (2.8%) |
| Missing | 193 (8.2%) | 119 (7.1%) |
| Abbreviation: COVID-19 = Coronavirus Disease of 2019; DMT<br>= disease-modifying therapy, EDSS = Expanded Disability<br>Status Scale; ICU = Intensive care unit; MS = multiple sclerosis;<br>RRMS = relapsing-remitting multiple sclerosis. |  |  |

### Cohort characteristics

Data sources were located in: 1) Sweden (n=290); 2) Australia, Belgium, Brasil, Kuwait, Romania, Saudi Arabia, Turkey (n=97); 3) Argentina, Chile, Colombia, Ecuador, Honduras, Mexico (n=159); 4) Bulgaria (n=3); 5) Germany, Italy (n=45); 6) Denmark (n=56); 7) Brasil (n=96); 8) Australia, Bahamas, Belgium, Czech Republic, Finland, France, Netherlands, New Zealand, Serbia, Spain, UK, USA (n=114); 9) Germany (n=41); 10) USA (n=1,161); 11) UK (n=131); 12) Spain (n=147). Sources-2 and 10 had higher proportions with confirmed COVID-19 and Sources-4 and 5 higher with non-suspected COVID-19. Among suspected/confirmed COVID-19 cases, hospitalisation was higher in Source-11 and lower in Sources-2, 4, 5, and 7; ICU admission was higher in Sources-3, 9, and 10, and lower in Sources-2, 4, 7, 9, 11, and 12; ventilation was higher in Sources-3 and 8, and lower in Sources-2, 4, 5, 6, 7, 9, and 11; death was higher in Source-11 and lower in Sources-2, 4, 5, 6, 7, 8, and 9. Results were comparable on restriction to confirmed-only COVID-19 (data not shown).

Compared to dimethyl fumarate, a lower proportion of females were untreated or treated with interferon or ocrelizumab (Table S1). Larger proportions of those aged 50-69 and ≥70 were untreated or treated with ocrelizumab, teriflunomide, or Other DMT. Greater proportions of progressive MS patients were untreated or treated with ocrelizumab, rituximab, or Other DMTs. Of more disabled patients (EDSS>6), higher proportions were either untreated or treated with ocrelizumab or Other DMTs. Similar results were seen among confirmed-only COVID-19 (data not shown).

## COVID-19 severity, by clinical and demographic characteristics

Demographic and clinical characteristics of hospitalisation and ICU admission (Table 2) and requiring artificial ventilation and death (Table 3) among suspected/confirmed COVID-19 cases were assessed. In multivariable models, female sex showed an inverse trend with risks of hospitalisation and death, while older age was positively associated with hospitalisation and death. Progressive MS-phenotype was associated with higher risk of hospitalisation. Higher EDSS was associated with higher risks of all outcomes. Among confirmed-only COVID-19, most of these associations persisted (Tables S2-S3).

**Table 2.**
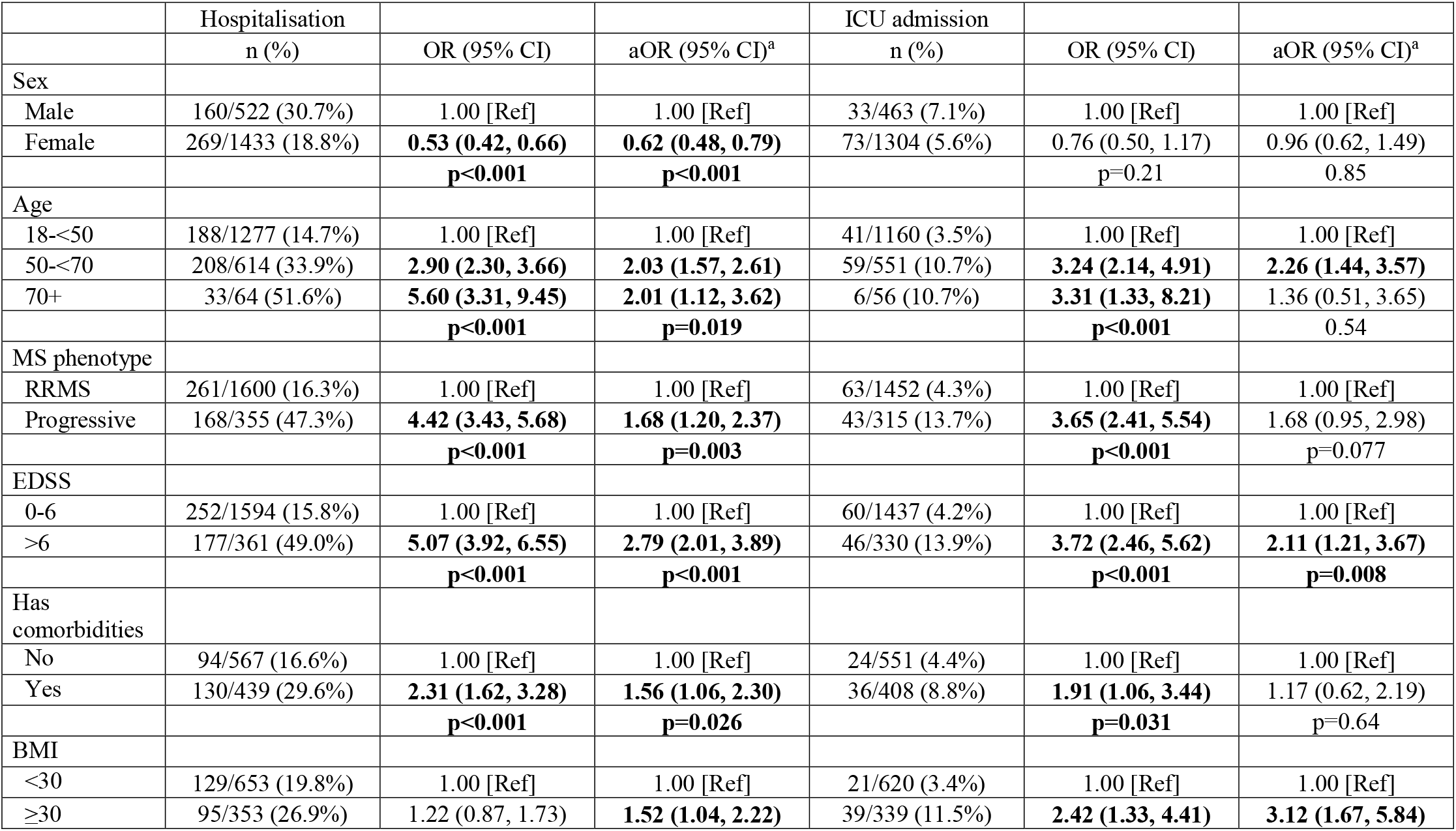
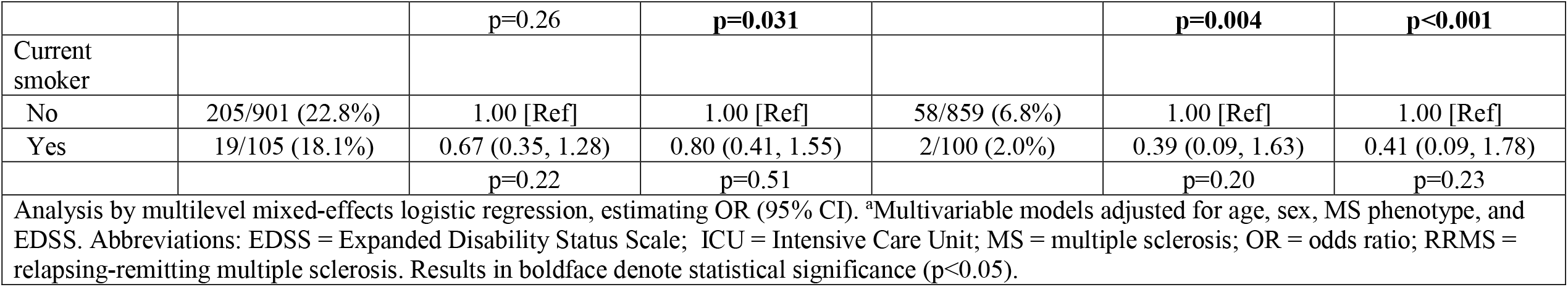
Demographic and clinical characteristics of hospitalisation & ICU admission, suspected+confirmed COVID-19.

**Table 3.**
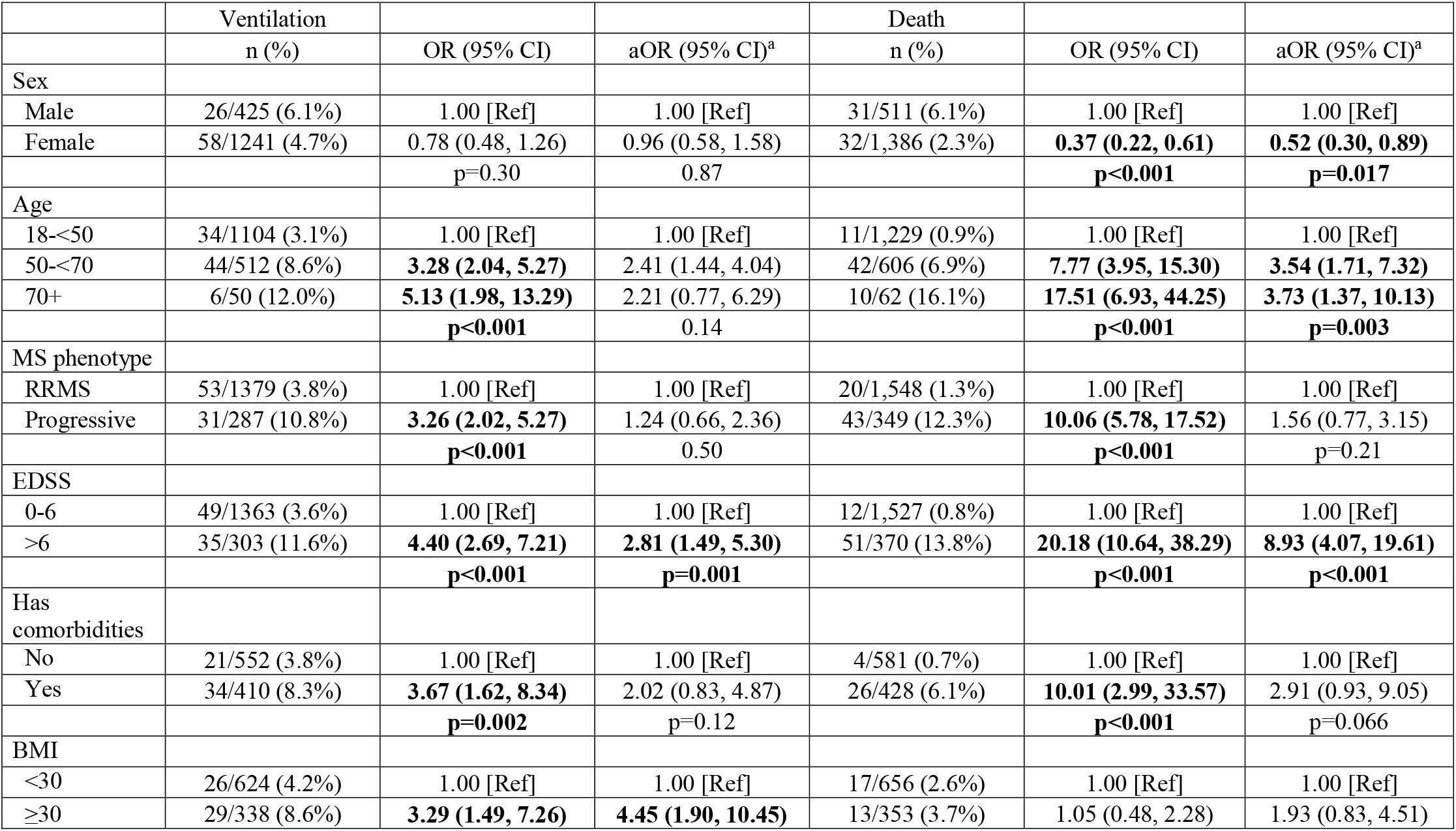
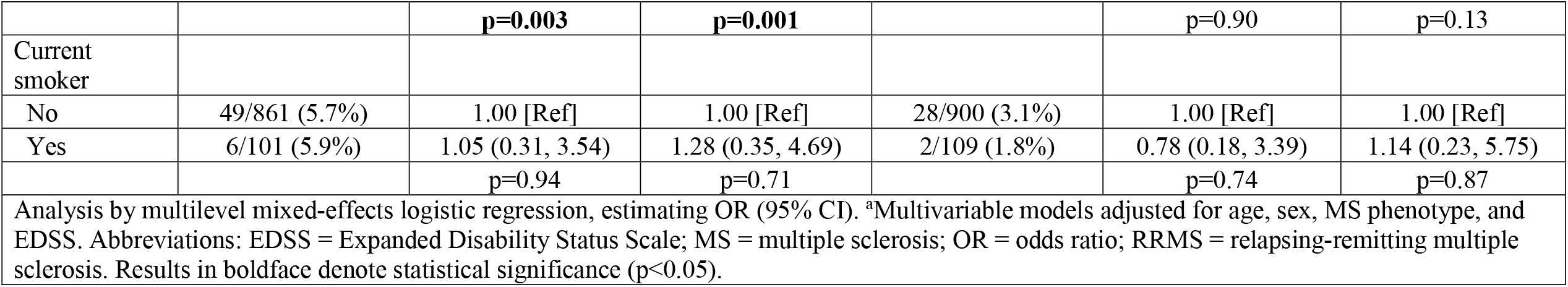
Demographic and clinical characteristics of ventilation & death, suspected+confirmed COVID-19.

In the subset of data sources with data available, having comorbidities was associated with increased risk of death and obese BMI with increased risks of hospitalisation, ICU admission, and ventilation, while smoking was not associated with any outcomes.

## COVID-19 severity, by DMT

Compared to dimethyl fumarate, rituximab use was associated with greater risks of hospitalisation (aOR=2.43), ICU admission (aOR=3.93), and artificial ventilation (aOR=4.00, Table 4). Ocrelizumab showed similar trends for hospitalisation (aOR=1.56) and ICU admission (aOR=2.30), but not artificial ventilation (aOR=1.04). No DMTs were associated with death. Untreated patients had increased risk of hospitalisation (aOR=1.79) but no independent associations with other outcomes were seen. These associations persisted among confirmed-only COVID-19 (Figures 1-3, Table S4).

**Table 4.** Characteristics of COVID-19 severity outcomes, DMTs vs dimethyl fumarate, anti-CD20 DMTs vs all other DMTs, and anti-CD20 DMTs vs natalizumab, suspected+confirmed COVID-19.

|  | Hospitalisation |  |  | ICU admission |  |  |
| --- | --- | --- | --- | --- | --- | --- |
|  | n (%) | OR (95% CI) | aOR (95% CI)<sup>a</sup> | n (%) | OR (95% CI) | aOR (95% CI)<sup>a</sup> |
| DMT |  |  |  |  |  |  |
| Untreated | 97/266 (36.5%) | **3.34 (2.17, 5.16)** | **1.79 (1.12, 2.87)** | 17/261 (6.5%) | **2.62 (1.06, 6.48)** | 1.36 (0.52, 3.55) |
| Alemtuzumab | 3/30 (10.0%) | 0.70 (0.20, 2.47) | 0.86 (0.24, 3.07) | 1/30 (3.3%) | 1.41 (0.16, 12.09) | 1.93 (0.22, 16.77) |
| Cladribine | 2/27 (7.4%) | 0.60 (0.13, 2.72) | 0.67 (0.14, 3.16) | 0/27 (0.0%) |  |  |
| Dimethyl fumarate | 38/268 (14.2%) | 1.00 [Ref] | 1.00 [Ref] | 7/259 (2.7%) | 1.00 [Ref] | 1.00 [Ref] |
| Fingolimod | 16/195 (8.2%) | 0.60 (0.32, 1.11) | 0.64 (0.34, 1.21) | 4/193 (2.1%) | 0.80 (0.23, 2.79) | 0.90 (0.25, 3.19) |
| Glatiramer acetate | 16/86 (18.6%) | 1.36 (0.71, 2.61) | 1.06 (0.53, 2.10) | 0/86 (0.0%) |  |  |
| Interferon | 17/119 (14.3%) | 1.11 (0.59, 2.11) | 0.89 (0.46, 1.70) | 2/118 (1.7%) | 0.62 (0.13, 3.10) | 0.54 (0.11, 2.73) |
| Natalizumab | 24/212 (11.3%) | 0.84 (0.48, 1.46) | 0.82 (0.47, 1.45) | 6/209 (2.9%) | 1.07 (0.35, 3.25) | 1.11 (0.36, 3.43) |
| Ocrelizumab | 116/463 (25.1%) | **2.14 (1.41, 3.24)** | **1.56 (1.01, 2.41)** | 37/463 (8.0%) | **3.00 (1.30, 6.89)** | 2.30 (0.98, 5.39) |
| Rituximab | 70/252 (27.8%) | **3.12 (1.92, 5.07)** | **2.43 (1.48, 4.02)** | 24/251 (9.6%) | **4.63 (1.86, 11.49)** | **3.93 (1.56, 9.89)** |
| Teriflunomide | 14/93 (15.1%) | 1.21 (0.61, 2.38) | 0.85 (0.42, 1.72) | 4/93 (4.3%) | 1.59 (0.45, 5.59) | 1.15 (0.32, 4.14) |
| Other DMT | 16/68 (23.5%) | **2.09 (1.07, 4.09)** | 1.10 (0.54, 2.24) | 4/67 (6.0%) | 2.34 (0.66, 8.33) | 1.27 (0.34, 4.72) |
| Pooled Other DMT | 144/1082 (13.3%) | 1.00 [Ref] | 1.00 [Ref] | 28/1082 (2.6%) | 1.00 [Ref] | 1.00 [Ref] |
| Ocrelizumab | 116/463 (25.1%) | **2.15 (1.62, 2.87)** | **1.75 (1.29, 2.38)** | 37/463 (8.0%) | **3.08 (1.83, 5.17)** | **2.55 (1.49, 4.36)** |
| Rituximab | 70/251 (27.9%) | **3.16 (2.17, 4.61)** | **2.76 (1.87, 4.07)** | 24/251 (9.6%) | **4.69 (2.49, 8.83)** | **4.32 (2.27, 8.23)** |
| No DMT | 97/261 (37.2%) | **3.49 (2.54, 4.80)** | **2.05 (1.43, 2.94)** | 17/261 (6.5%) | **2.69 (1.43, 5.04)** | 1.52 (0.77, 3.02) |
| Natalizumab | 24/209 (11.5%) | 1.00 [Ref] | 1.00 [Ref] | 6/209 (2.9%) | 1.00 [Ref] | 1.00 [Ref] |
| Ocrelizumab | 116/463 (25.1%) | **2.64 (1.64, 4.25)** | **1.86 (1.13, 3.07)** | 37/463 (8.0%) | **2.89 (1.18, 7.05)** | 2.13 (0.85, 5.35) |
| Rituximab | 70/251 (27.9%) | **3.16 (1.89, 5.26)** | **2.88 (1.68, 4.92)** | 24/251 (9.6%) | **4.17 (1.55, 11.18)** | **3.23 (1.17, 8.91)** |

|  | Ventilation |  |  | Death |  |  |
| --- | --- | --- | --- | --- | --- | --- |
|  | n (%) | OR (95% CI) | aOR (95% CI)<sup>a</sup> | n (%) | OR (95% CI) | aOR (95% CI)<sup>a</sup> |
| DMT |  |  |  |  |  |  |
| Untreated | 18/261 (6.9%) | **2.51 (1.00, 6.26)** | 1.31 (0.49, 3.49) | 27/261 (10.3%) | **5.69 (2.13, 15.19)** | 1.64 (0.56, 4.80) |
| Alemtuzumab | 1/30 (3.3%) | 0.98 (0.11, 8.63) | 1.27 (0.14, 11.34) | 1/30 (3.3%) | 2.10 (0.23, 19.15) | 2.11 (0.19, 23.29) |
| Cladribine | 0/27 (0.0%) |  |  | 0/27 (0.0%) |  |  |
| Dimethyl fumarate | 7/259 (2.7%) | 1.00 [Ref] | 1.00 [Ref] | 5/259 (1.9%) | 1.00 [Ref] | 1.00 [Ref] |
| Fingolimod | 5/193 (2.6%) | 0.76 (0.23, 2.50) | 0.79 (0.24, 2.67) | 0/193 (0.0%) |  |  |
| Glatiramer acetate | 0/86 (0.0%) |  |  | 2/86 (2.3%) | 1.05 (0.20, 5.57) | 0.56 (0.10, 3.25) |
| Interferon | 1/118 (0.8%) | 0.21 (0.02, 1.75) | 0.18 (0.02, 1.52) | 1/118 (0.8%) | 0.58 (0.07, 5.20) | 0.31 (0.03, 2.82) |
| Natalizumab | 4/209 (1.9%) | 0.74 (0.21, 2.59) | 0.72 (0.20, 2.55) | 3/209 (1.4%) | 0.78 (0.18, 3.33) | 0.72 (0.16, 3.30) |
| Ocrelizumab | 18/463 (3.9%) | 1.48 (0.60, 3.66) | 1.04 (0.41, 2.64) | 12/463 (2.6%) | 1.22 (0.42, 3.56) | 0.49 (0.16, 1.52) |
| Rituximab | 24/251 (9.6%) | **5.11 (2.01, 12.99)** | **4.00 (1.54, 10.39)** | 7/251 (2.8%) | 2.61 (0.76, 8.96) | 1.22 (0.35, 4.18) |
| Teriflunomide | 4/93 (4.3%) | 1.33 (0.37, 4.79) | 0.83 (0.22, 3.10) | 1/93 (1.1%) | 0.59 (0.07, 5.21) | 0.27 (0.03, 2.53) |
| Other DMT | 2/67 (3.0%) | 1.12 (0.22, 5.71) | 0.61 (0.12, 3.21) | 4/67 (6.0%) | 3.82 (0.97, 15.00) | 1.05 (0.25, 4.45) |
| Pooled Other DMT | 24/1082 (2.2%) | 1.00 [Ref] | 1.00 [Ref] | 17/1082 (1.6%) | 1.00 [Ref] | 1.00 [Ref] |
| Ocrelizumab | 18/463 (3.9%) | **1.99 (1.04, 3.80)** | 1.60 (0.82, 3.14) | 12/463 (2.6%) | 1.38 (0.65, 2.96) | 0.76 (0.34, 1.69) |
| Rituximab | 24/251 (9.6%) | **6.95 (3.54, 13.64)** | **6.15 (3.09, 12.27)** | 7/251 (2.8%) | **2.94 (1.12, 7.72)** | 1.90 (0.73, 4.93) |
| No DMT | 18/261 (6.9%) | **3.42 (1.78, 6.57)** | **2.07 (1.01, 4.22)** | 27/261 (10.3%) | **6.47 (3.40, 12.33)** | **2.53 (1.24, 5.15)** |
| Natalizumab | 4/209 (1.9%) | 1.00 [Ref] | 1.00 [Ref] | 3/209 (1.4%) | 1.00 [Ref] | 1.00 [Ref] |
| Ocrelizumab | 18/463 (3.9%) | 2.11 (0.69, 6.39) | 1.34 (0.42, 4.24) | 12/463 (2.6%) | 1.64 (0.45, 5.96) | 0.53 (0.13, 2.24) |
| Rituximab | 24/251 (9.6%) | **7.52 (2.37, 23.81)** | **5.52 (1.71, 17.84)** | 7/251 (2.8%) | 2.96 (0.66, 13.29) | 1.70 (0.38, 7.62) |

Analysis by multilevel mixed-effects logistic regression, estimating OR (95% CI). <sup>a</sup>Multivariable models adjusted for age, sex, MS phenotype, and EDSS. Abbreviations: DMT = disease-modifying therapy; EDSS = Expanded Disability Status Scale; ICU = Intensive Care Unit; MS = multiple sclerosis; OR = odds ratio. Results in boldface denote statistical significance (p<0.05).

**Figure 1.**
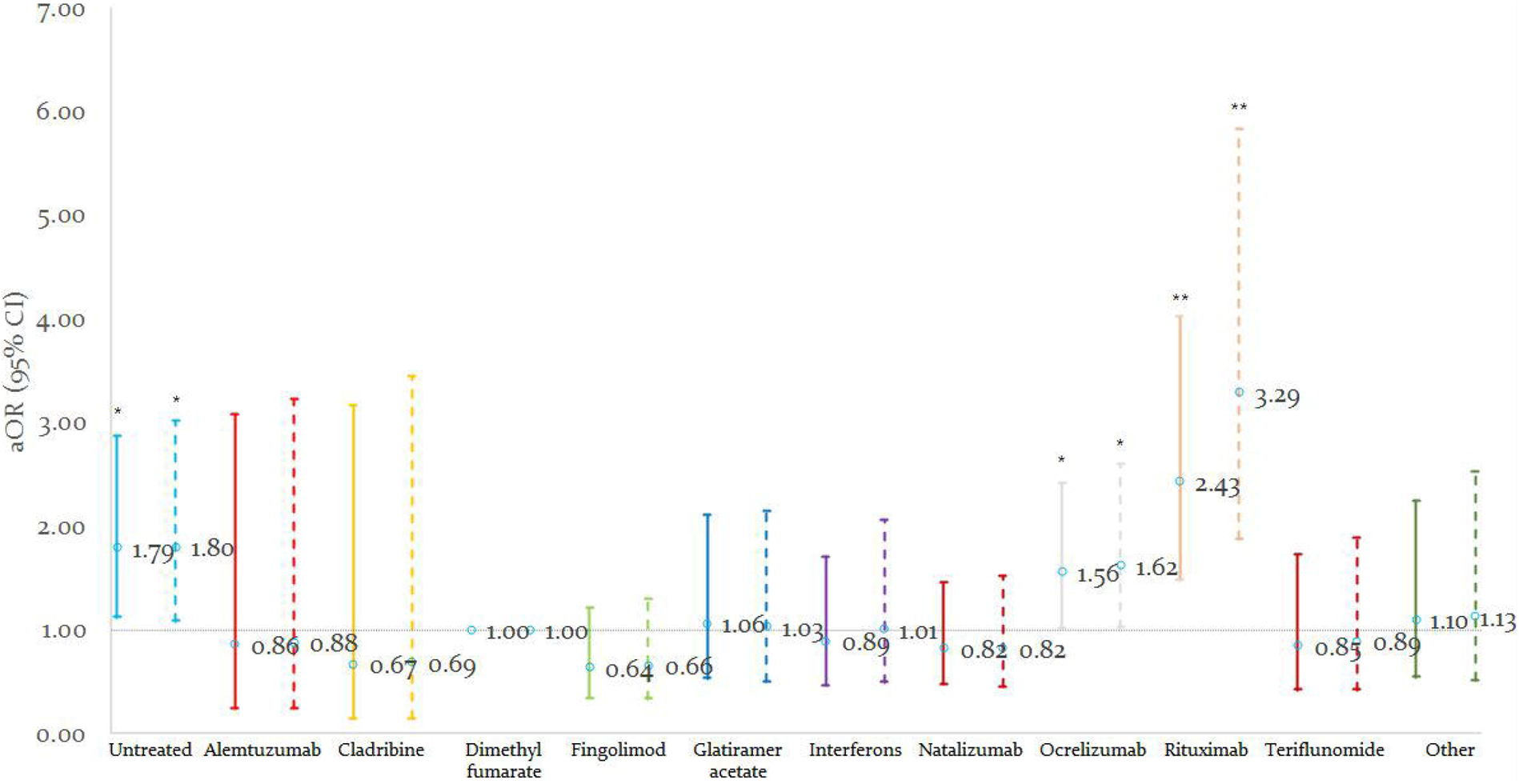
The risk of hospitalisation by DMT among suspected/confirmed (solid line) and confirmed-only (dashed line) cases. DMTs compared to dimethyl fumarate, adjusted for age, sex, MS-phenotype, and EDSS. Note: Other DMTs also includes siponimod. * = p<0.05, ** = p<0.001.

**Figure 2:**
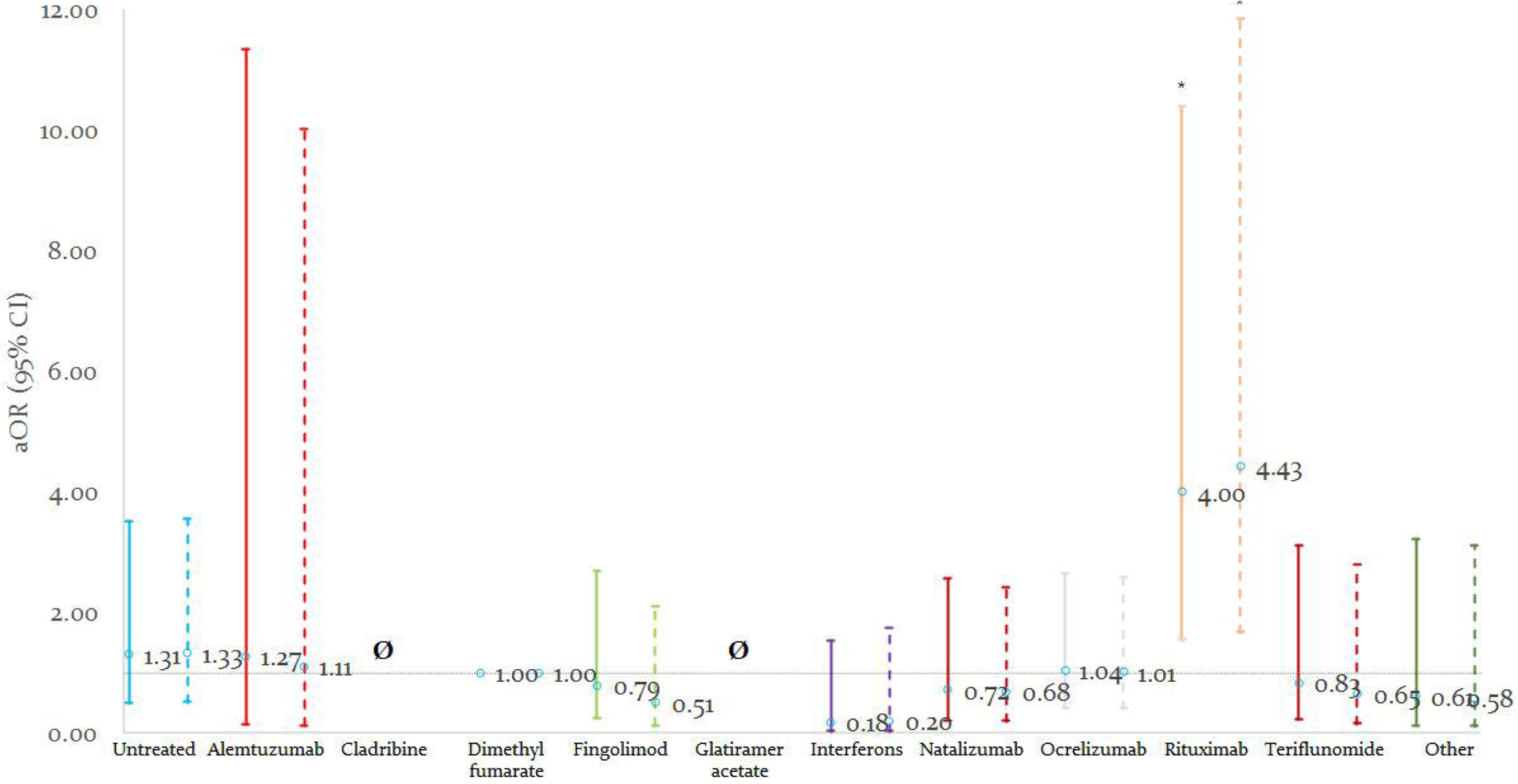
The risk of ICU admission by DMT, among suspected/confirmed (solid line) and confirmed-only (dashed line) cases. DMTs were compared to dimethyl fumarate, adjusted for age, sex, MS-phenotype, and EDSS. Note: Other DMTs also includes siponimod. * = p<0.05. Note: null set denotes analyses which could not be undertaken due to no events occurring in the exposed group.

**Figure 3:**
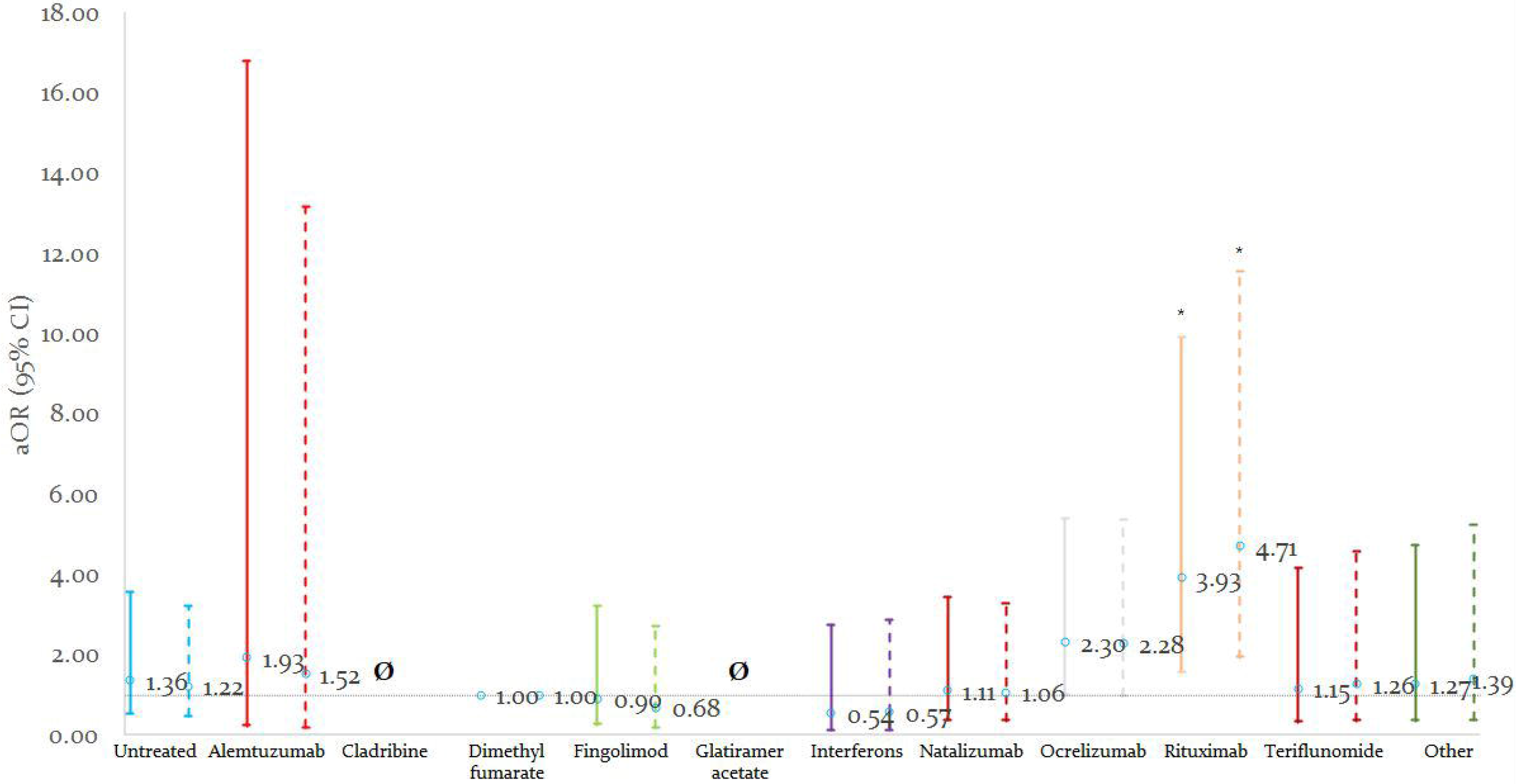
The risk of artificial ventilation by DMT, among suspected/confirmed (solid line) and confirmed-only (dashed line) cases. DMTs were compared to dimethyl fumarate, adjusted for age, sex, MS-phenotype, and EDSS. Note: Other DMTs also includes siponimod. * = p<0.05. Note: null set denotes analyses which could not be undertaken due to no events occurring in the exposed group.

## COVID-19 severity: anti-CD20 DMTs vs pooled other DMTs

Compared to all other DMTs (Table 4), those using rituximab had higher risks of hospitalisation (aOR=2.76), ICU admission (aOR=4.32), and artificial ventilation (aOR=6.15). Ocrelizumab showed similar trends for hospitalisation (aOR=1.75) and ICU admission (aOR=2.55) but not ventilation (aOR=1.60). Neither rituximab or ocrelizumab were associated with risk of death. Untreated patients had increased risks of hospitalisation (aOR=2.05), ventilation (aOR=2.07), and death (aOR=2.53). These results persisted among confirmed-only COVID-19 cases (Table S5).

## COVID-19 severity, anti-CD20 DMTs vs natalizumab

Compared to natalizumab, rituximab was associated with higher risks of hospitalisation (aOR=2.88), ICU admission (aOR=3.23), and ventilation (aOR=5.52, Table 4). Ocrelizumab showed similar trends for hospitalisation (aOR=1.86), but did not reach significance for ICU admission and was not associated with ventilation. Neither rituximab or ocrelizumab was associated with increased risk of death. Results were similar among confirmed-only COVID-19 (Table S5).

## Mitigating heterogeneity among data sources

To mitigate the impact of potential heterogeneity among the individual data sources influencing the results, particularly that for source C10 which comprised nearly half the sample, we utilised random-effects logistic regression. However, in addition, serial exclusion analyses were undertaken wherein analyses were run excluding each dataset in turn. These analyses showed no material differences in associations compared to the main analysis (data not shown).

## Stratification sensitivity analyses

To assess whether the associations seen for ocrelizumab and rituximab with outcomes was genuinely a function of the DMTs, rather than the characteristics of patients commonly treated with these medications (older, progressive MS, higher disability), we undertook stratified analyses evaluating associations among persons aged>70 years vs ≤70, among RRMS vs progressive, and among EDSS≤6 vs EDSS>6. These analyses showed that associations were actually present in persons of younger age, of RRMS phenotype, and lower EDSS, indicating that the observed associations were a function of the DMT, not underlying risk profile (Tables S6-S8).

## Discussion

In the largest sample of people with MS with suspected and confirmed COVID-19 to date, we demonstrated that the anti-CD20 DMTs rituximab, and to a lesser extent ocrelizumab, were associated with more severe COVID-19. Compared to dimethyl fumarate, pooled other DMTs, and natalizumab, anti-CD20 DMTs were associated with higher risks of hospitalisation and ICU admission, while only rituximab was associated with greater risk of requiring artificial ventilation for each analysis. Comparison to natalizumab is particularly important, suggesting that anti-CD20 associations do not reflect ascertainment bias. Regardless of comparator, rituximab consistently showed stronger associations with outcomes than ocrelizumab for hospitalisation and ICU admission, and was solely associated with requiring artificial ventilation. Moreover, DMT associations were not merely driven by older patients, progressive MS-phenotype, or higher disability. We also found that older age, progressive MS-phenotype, and higher disability were over-represented among MS patients with more severe COVID-19. In sub-analyses where data was available, DMT associations were robust to further adjustment for comorbidities, BMI, and smoking status.

Anti-CD20 DMTs, which selectively deplete circulating B-lymphocytes, alemtuzumab and cladribine which act through broader immunosuppressive mechanisms, and fingolimod, which sequesters lymphocytes from circulation, are highly effective MS treatments, but can increase infection risk^1, 16^. This has raised concern during the COVID-19 pandemic and national studies have investigated risk factors for severe COVID-19 disease in people with MS. In the French COVISEP study^7^ of 347 MS patients (42.1% confirmed COVID-19), older age, progressive MS-phenotype, higher disability, and comorbidities were associated with COVID-19 severity. In addition, pooled DMTs with moderate/high risk of systemic infection (fingolimod, ocrelizumab, rituximab, cladribine, alemtuzumab) were associated with 4.2-times higher COVID-19 severity score than DMTs with no systemic infection risk (interferon-□, glatiramer-acetate). This amalgamation of DMTs is a limitation since, while comparable in terms of their infection risk, these DMTs have markedly different modes of action, especially in relation to the immunological response to SARS-CoV-2^15^. More recently, the Italian MuSC-19 national registry study of 593 suspected and 191 confirmed COVID-19 cases, found anti-CD20 DMT use was associated with 2.6-times greater risk of severe COVID-19 compared to dimethyl fumarate, adjusted for region, age, sex, MS-phenotype, and recent methylprednisolone use^9^.

Untreated patients showed consistent positive trends towards associations with hospitalisation, ICU admission, and requiring ventilation, albeit attenuating on adjustment. This is in keeping with prior results. Louapre and colleagues found higher frequencies of severe COVID-19 among the untreated vs treated (46.0% vs 15.5%), though this difference did not persist on adjustment^7^. Sormani and colleagues compared untreated to dimethyl fumarate-treated, finding they were 2.83-times more likely to have severe COVID-19, though this disappeared on adjustment (aOR=1.04)^9^. The lack of independence of the untreated associations here and previously likely reflects the untreated comprising to variable degree people with more benign MS course or other reasons to not use DMTs, so adjustment for MS-phenotype and disability largely captures differences in COVID-19 severity.

Of interest are the differences in the magnitudes of associations seen for ocrelizumab and rituximab, since both bind the same CD20 epitope on the outer loop of the protein, albeit in slightly different fashions^17, 18^. In comparison to dimethyl fumarate, to pooled other DMTs, and to natalizumab, rituximab consistently showed stronger associations with outcomes than ocrelizumab. Importantly, in analyses stratifying associations of DMT exposure with COVID19 severity by age, MS-phenotype, and EDSS, there was no evidence that the anti-CD20 DMT associations merely reflects patients with underlying risk factors being disposed to use these treatments. The differences seen, both in magnitudes of associations with hospitalisation and ICU admission and the lack of association with ventilation, are puzzling. This weaker magnitude for ocrelizumab has been seen in prior iterations of the analysis but also in those prior iterations ocrelizumab was significantly associated with ventilation as well as hospitalisation and ICU admission, albeit of weaker magnitude^11^. That ocrelizumab was no longer associated in this iteration of the data may reflect changes in medical practice for COVID-19 treatment, including the introduction of more effective treatments that reduce the need for patients to require artificial ventilation. The weaker magnitudes of associations may reflect ocrelizumab’s slightly lower affinity to CD20 or possibly their different provenance, rituximab being a chimeric IgG while ocrelizumab is humanised^17, 18^. Also, the mode of B-cell elimination differs, with ocrelizumab using more antibody and less complement-mediated cytotoxicity compared to rituximab^19^. Alternatively, this difference may represent unmeasured confounding as the dataset, while large, was limited in the number of characteristics assessed, so potentially relevant factors like socioeconomic status and access to care or factors impacting on respiratory health could not be assessed. That said, our results are broadly in line with those seen in other studies^7, 9^, providing external consistency. This preliminary observation is worth exploring in laboratory studies.

That anti-CD20 DMTs were not associated with death conflicts with the results seen for the other outcomes, as well as with the MuSC-19 study which found a positive trend between anti-CD20 DMTs and death. The issue may lie in ascertainment bias, with fewer of the older patients included in our sample: we had only 9.1% of confirmed COVID-19 cases over 60 years old, vs 17.7% in the MuSC-19 cohort^9^. The potential impacts of these DMTs on death due to COVID-19 should be further explored.

### Limitations

In contrast to prior clinic-based studies, our cohort focused on a pre-defined limited set of demographic and clinical characteristics^10^. Thus, we could not assess other clinical features, particularly prior MS clinical course and DMT use, paraclinical information such as radiological burden of MS, or the nuanced details of COVID-19 onset and evolution. Another limitation of our data is that they likely comprise greater proportions of severe cases requiring medical attention. One particular element lacking in our data is treatment duration or duration since treatment, since these both may have bearing on the degree of B-cell depletion and thence on COVID-19 severity. This data was included in the core questionnaire but the level of missingness was too high to be a component of analyses.

Heterogeneity in the definitions of exposure and outcomes and in patient inclusion among the data-sources is a known problem in combining multiple data-sources. Related to this are the differences in protocols for hospital and ICU admission and initiation of artificial ventilation between hospitals. To ensure that our results were not being driven by single influential data-sources, we undertook all analyses using random-effects logistic regression, as well as serial-exclusion sensitivity analyses. These analyses showed that, while there was some variation in the magnitudes and significance of associations, trends tracked as seen for the whole cohort, indicating that the results were not driven by a specific data-source.

Another issue lies in the anonymous nature of the data entry, such that patients may be entered more than once in different data-sources. We are unable to account for whether participants already participated or had their data entered in another study as there is no identifying information to assess this, nor any query of prior participation in the survey.

Another issue is the comparator used for individual DMTs. We initially planned to compare to glatiramer acetate given its absence of impact on infection risk; however, infrequency of outcomes among these patients precluded its being the comparator. Interferon-beta and teriflunomide were potential comparators but their potential impacts on infection immune response argued against this. Dimethyl fumarate was identified as a suitable comparator, being common in the sample and was also that used for the MuSC-19 study^9^. The untreated were not regarded as an appropriate primary comparator because these patients differed markedly from the rest of the cohort in age, disability, and MS-phenotype.

Another issue is the nature of the data aggregation, with some data sources providing individual patient-level data but others only tabulations of discrete categorical terms. Thus, we were obliged to use three-level categories of age and two-level EDSS, rather than more exact values of each. That said, these levels are generally aligned with the levels of each associated with increased COVID-19 severity.

Another issue is the definition of outcome values. Where these values were missing, we have left them as such, rather than inferring for instance that patients who were not hospitalised were also not admitted to ICU or required artificial ventilation. This was a conservative allocation that increased the proportions missing for such parameters, rather than coding them to null values, but analyses coding such to null values made little impact on results (data not shown).

### Conclusions

In the largest population yet studied, we have shown that MS patients treated with anti-CD20 DMTs, rituximab and ocrelizumab, are at higher risk of more severe COVID-19 compared to those treated with dimethyl fumarate, to pooled other DMTs, and to natalizumab. This risk is additional to the risk associated with demographic and clinical characteristics. These results agree with smaller cohort studies and suggest that the risk-vs-benefit of continued or new exposure to CD20-depleting treatment strategies compared to other DMTs needs to be considered in the context of the ongoing COVID-19 pandemic.

## Supporting information

STROBE checklist

## Data Availability

Persons interested in acquiring the data used for this analysis may contact Professor Dr Liesbet Peeters in regards.

