## Supplemental Tables 1-8 for "Associations of DMT therapies with COVID-19 severity in multiple sclerosis"

Table S1. Clinical and demographic characteristics of DMT use, suspected+confirmed COVID-19.

|  | Untreated (n=276) | Amab (n=31) | Cladribine (n=29) | Dimethyl fumarate (n=264) | Fingolimod (n=197) | Glatiramer acetate (n=87) | Interferon (n=121) | Nmab (n=215) | Omab (n=471) | Rmab (n=257) | Terf (n=97) | Other DMT (n=69) |
| --- | --- | --- | --- | --- | --- | --- | --- | --- | --- | --- | --- | --- |
| Sex |  |  |  |  |  |  |  |  |  |  |  |  |
| Male | 81 (29.3%) | 7 (22.6%) | 6 (20.7%) | 63 (23.9%) | 46 (23.4%) | 19 (21.8%) | 36 (29.8%) | 44 (20.5%) | 153 (32.5%) | 70 (27.2%) | 28 (28.9%) | 15 (21.7%) |
| Female | 195 (70.7%) | 24 (77.4%) | 23 (79.3%) | 201 (76.1%) | 151 (76.6%) | 68 (78.2%) | 84 (69.4%) | 171 (79.5%) | 318 (67.5%) | 187 (72.8%) | 69 (71.1%) | 54 (78.3%) |
| Missing | 0 (0.0%) | 0 (0.0%) | 0 (0.0%) | 0 (0.0%) | 0 (0.0%) | 0 (0.0%) | 1 (0.8%) | 0 (0.0%) | 0 (0.0%) | 0 (0.0%) | 0 (0.0%) | 0 (0.0%) |
| Age |  |  |  |  |  |  |  |  |  |  |  |  |
| 18-<50 | 121 (43.8%) | 26 (83.9%) | 22 (75.9%) | 183 (69.3%) | 155 (78.7%) | 55 (63.2%) | 74 (61.2%) | 168 (78.1%) | 300 (63.7%) | 187 (72.8%) | 46 (47.4%) | 39 (56.5%) |
| 50-<70 | 118 (42.8%) | 4 (12.9%) | 5 (17.2%) | 79 (29.9%) | 40 (20.3%) | 26 (29.9%) | 44 (36.4%) | 43 (20.0%) | 161 (34.2%) | 67 (26.1%) | 48 (49.5%) | 26 (37.7%) |
| 70+ | 37 (13.4%) | 0 (0.0%) | 0 (0.0%) | 2 (0.8%) | 0 (0.0%) | 6 (6.9%) | 2 (1.7%) | 3 (1.4%) | 7 (1.5%) | 1 (0.4%) | 3 (3.1%) | 4 (5.8%) |
| Missing | 0 (0.0%) | 1 (3.2%) | 2 (6.9%) | 0 (0.0%) | 2 (1.0%) | 0 (0.0%) | 1 (0.8%) | 1 (0.5%) | 3 (0.6%) | 2 (0.8%) | 0 (0.0%) | 0 (0.0%) |
| MS phenotype |  |  |  |  |  |  |  |  |  |  |  |  |
| RRMS | 152 (55.1%) | 30 (96.8%) | 28 (96.6%) | 242 (91.7%) | 188 (95.4%) | 74 (85.1%) | 104 (86.0%) | 204 (94.9%) | 376 (79.8%) | 200 (77.8%) | 90 (92.8%) | 46 (66.7%) |
| Progressive | 124 (44.9%) | 1 (3.2%) | 1 (3.4%) | 21 (8.0%) | 9 (4.6%) | 13 (14.9%) | 17 (14.0%) | 11 (5.1%) | 95 (20.2%) | 56 (21.8%) | 7 (7.2%) | 22 (31.9%) |
| Missing | 0 (0.0%) | 0 (0.0%) | 0 (0.0%) | 1 (0.4%) | 0 (0.0%) | 0 (0.0%) | 0 (0.0%) | 0 (0.0%) | 0 (0.0%) | 1 (0.4%) | 0 (0.0%) | 1 (1.4%) |
| EDSS |  |  |  |  |  |  |  |  |  |  |  |  |
| 0-6 | 162 (58.7%) | 29 (93.5%) | 27 (93.1%) | 239 (90.5%) | 187 (94.9%) | 69 (79.3%) | 111 (91.7%) | 183 (85.1%) | 339 (72.0%) | 213 (82.9%) | 81 (83.5%) | 43 (62.3%) |
| >6 | 99 (35.9%) | 2 (6.5%) | 2 (6.9%) | 20 (7.6%) | 8 (4.1%) | 17 (19.5%) | 9 (7.4%) | 27 (12.6%) | 127 (27.0%) | 40 (15.6%) | 12 (12.4%) | 24 (34.8%) |
| Missing | 15 (5.4%) | 0 (0.0%) | 0 (0.0%) | 5 (1.9%) | 2 (1.0%) | 1 (1.1%) | 1 (0.8%) | 5 (2.3%) | 5 (1.1%) | 4 (1.6%) | 4 (4.1%) | 2 (2.9%) |
| Abbreviation: Amab = Alemtuzumab; DMT = disease-modifying therapy, EDSS = Expanded Disability Status Scale; ICU = Intensive care unit; MS = multiple sclerosis; Nmab = Natalizumab; Rmab = Rituximab; RRMS = relapsing-remitting multiple sclerosis; Terf = Teriflunomide. | | | | | | | | | | | | |

Table S2. Clinician-reported demographic and clinical characteristics of hospital & ICU admission, confirmed COVID-19 only.

|  | Hospitalisation |  |  | ICU admission |  |  |
| --- | --- | --- | --- | --- | --- | --- |
|  | n (%) | OR (95% CI) | aOR (95% CI)^a^ | n (%) | OR (95% CI) | aOR (95% CI)^a^ |
| Sex |  |  |  |  |  |  |
| Male | 147/391 (37.6%) | 1.00 [Ref] | 1.00 [Ref] | 33/357 (9.2%) | 1.00 [Ref] | 1.00 [Ref] |
| Female | 250/1050 (23.8%) | **0.53 (0.41, 0.68)** | **0.64 (0.49, 0.84)** | 70/982 (7.1%) | 0.75 (0.48, 1.16) | 0.96 (0.61, 1.50) |
|  |  | **p<0.001** | **p=0.001** |  | p=0.19 | p=0.84 |
| Age |  |  |  |  |  |  |
| 18-<50 | 172/900 (19.1%) | 1.00 [Ref] | 1.00 [Ref] | 40/844 (4.7%) | 1.00 [Ref] | 1.00 [Ref] |
| 50-<70 | 192/484 (39.7%) | 4/136 (2.9%) | **1.85 (1.40, 2.44)** | 57/446 (12.8%) | **2.95 (1.93, 4.49)** | **2.05 (1.29, 3.27)** |
| 70+ | 33/57 (57.9%) | 18/305 (5.9%) | 1.85 (0.98, 3.49) | 6/49 (12.2%) | **2.80 (1.13, 6.98)** | 1.28 (0.47, 3.45) |
|  |  | 23/107 (21.5%) | p=0.056 |  | **p<0.001** | p=0.63 |
| MS phenotype |  |  |  |  |  |  |
| RRMS | 238/1159 (20.5%) | 1.00 [Ref] | 1.00 [Ref] | 61/1083 (5.6%) | 1.00 [Ref] | 1.00 [Ref] |
| Progressive | 159/282 (56.4%) | **4.72 (3.56, 6.26)** | **1.94 (1.33, 2.84)** | 42/256 (16.4%) | **3.41 (2.22, 5.22)** | 1.68 (0.93, 3.04) |
|  |  | **p<0.001** | **p<0.001** |  | **p<0.001** | p=0.085 |
| EDSS |  |  |  |  |  |  |
| 0-6 | 229/1133 (20.2%) | 1.00 [Ref] | 1.00 [Ref] | 58/1058 (5.5%) | 1.00 [Ref] | 1.00 [Ref] |
| >6 | 168/308 (54.5%) | **4.72 (3.57, 6.23)** | **2.48 (1.73, 3.55)** | 45/281 (16.0%) | **3.34 (2.18, 5.10)** | **1.96 (1.11, 3.46)** |
|  |  | **p<0.001** | **p<0.001** |  | **p<0.001** | **p=0.020** |
| Has comorbidities |  |  |  |  |  |  |
| No | 91/469 (19.4%) | 1.00 [Ref] | 1.00 [Ref] | 24/454 (5.3%) | 1.00 [Ref] | 1.00 [Ref] |
| Yes | 120/375 (32.0%) | **2.03 (1.46, 2.82)** | **1.52 (1.08, 2.14)** | 35/345 (10.1%) | **2.02 (1.18, 3.47)** | 1.16 (0.65, 2.07) |
|  |  | **p<0.001** | **p=0.015** |  | **p=0.010** | p=0.61 |
| BMI |  |  |  |  |  |  |
| <30 | 122/538 (22.7%) | 1.00 [Ref] | 1.00 [Ref] | 21/505 (4.2%) | 1.00 [Ref] | 1.00 [Ref] |
| ≥30 | 89/306 (29.1%) | 1.28 (0.92, 1.79) | **1.53 (1.09, 2.14)** | 38/294 (12.9%) | **3.42 (1.97, 5.95)** | **3.78 (2.14, 6.66)** |
|  |  | p=0.14 | **p=0.014** |  | **p<0.001** | **p<0.001** |
| Current smoker |  |  |  |  |  |  |
| No | 195/766 (25.5%) | 1.00 [Ref] | 1.00 [Ref] | 57/725 (7.9%) | 1.00 [Ref] | 1.00 [Ref] |
| Yes | 16/78 (20.5%) | 0.74 (0.41, 1.31) | 0.83 (0.46, 1.50) | 2/74 (2.7%) | 0.32 (0.08, 1.35) | 0.36 (0.08, 1.57) |
|  |  | p=0.30 | p=0.53 |  | p=0.12 | p=0.17 |
| Analysis by multilevel mixed-effects logistic regression, estimating OR (95% CI). ^a^Multivariable models adjusted for age, sex, MS phenotype, and EDSS. Abbreviations: EDSS = Expanded Disability Status Scale; ICU = Intensive Care Unit; MS = multiple sclerosis; OR = odds ratio; RRMS = relapsing-remitting multiple sclerosis. Results in boldface denote statistical significance (p<0.05). | | | | | | |

Table S3. Clinician-reported demographic and clinical characteristics of ventilation & death, confirmed COVID-19 only.

|  | Ventilation |  |  | Death |  |  |
| --- | --- | --- | --- | --- | --- | --- |
|  | n (%) | OR (95% CI) | aOR (95% CI)^a^ | n (%) | OR (95% CI) | aOR (95% CI)^a^ |
| Sex |  |  |  |  |  |  |
| Male | 25/329 (7.6%) | 1.00 [Ref] | 1.00 [Ref] | 27/391 (6.9%) | 1.00 [Ref] | 1.00 [Ref] |
| Female | 55/948 (5.8%) | 0.79 (0.48, 1.30) | 1.01 (0.60, 1.70) | 28/1019 (2.8%) | **0.38 (0.22, 0.65)** | **0.56 (0.31, 1.00)** |
|  |  | p=0.35 | p=0.97 |  | **p<0.001** | **p=0.049** |
| Age |  |  |  |  |  |  |
| 18-<50 | 31/815 (3.8%) | 1.00 [Ref] | 1.00 [Ref] | 9/871 (1.0%) | 1.00 [Ref] | 1.00 [Ref] |
| 50-<70 | 43/418 (10.3%) | **3.10 (1.89, 5.10)** | **2.31 (1.35, 3.94)** | 36/484 (7.4%) | **7.62 (3.62, 16.03)** | **3.37 (1.52, 7.49)** |
| 70+ | 6/44 (13.6%) | **4.57 (1.71, 12.26)** | 2.10 (0.72, 6.19) | 10/55 (18.2%) | **19.71 (7.25, 53.60)** | **4.12 (1.43, 11.84)** |
|  |  | **p<0.001** | p=0.18 |  | **p<0.001** | **p=0.004** |
| MS phenotype |  |  |  |  |  |  |
| RRMS | 49/1044 (4.7%) | 1.00 [Ref] | 1.00 [Ref] | 16/1126 (1.4%) | 1.00 [Ref] | 1.00 [Ref] |
| Progressive | 31/233 (13.3%) | **3.31 (2.02, 5.44)** | 1.38 (0.71, 2.69) | 39/284 (13.7%) | **10.82 (5.89, 19.85)** | 1.97 (0.90, 4.28) |
|  |  | **p<0.001** | p=0.34 |  | **p<0.001** | p=0.088 |
| EDSS |  |  |  |  |  |  |
| 0-6 | 46/1017 (4.5%) | 1.00 [Ref] | 1.00 [Ref] | 11/1095 (1.0%) | 1.00 [Ref] | 1.00 [Ref] |
| >6 | 34/260 (13.1%) | **3.92 (2.37, 6.48)** | **2.45 (1.27, 4.73)** | 44/315 (14.0%) | **16.00 (8.15, 31.39)** | **6.11 (2.62, 14.21)** |
|  |  | **p<0.001** | **p=0.008** |  | **p<0.001** | **p<0.001** |
| Has comorbidities |  |  |  |  |  |  |
| No | 19/456 (4.2%) | 1.00 [Ref] | 1.00 [Ref] | 4/483 (0.8%) | 1.00 [Ref] | 1.00 [Ref] |
| Yes | 33/346 (9.5%) | **3.40 (1.77, 6.51)** | **2.21 (1.12, 4.33)** | 23/366 (6.3%) | **8.03 (2.75, 23.43)** | 2.77 (0.97, 7.89) |
|  |  | **p<0.001** | **p=0.021** |  | **p<0.001** | p=0.057 |
| BMI |  |  |  |  |  |  |
| <30 | 23/509 (4.5%) | 1.00 [Ref] | 1.00 [Ref] | 14/542 (2.6%) | 1.00 [Ref] | 1.00 [Ref] |
| ≥30 | 29/293 (9.9%) | **3.38 (1.78, 6.41)** | **3.14 (1.66, 5.93)** | 13/307 (4.2%) | 1.67 (0.77, 3.60) | **3.02 (1.32, 6.94)** |
|  |  | **p<0.001** | **p<0.001** |  | p=0.19 | **p=0.009** |
| Current smoker |  |  |  |  |  |  |
| No | 49/727 (6.7%) | 1.00 [Ref] | 1.00 [Ref] | 26/770 (3.4%) | 1.00 [Ref] | 1.00 [Ref] |
| Yes | 3/75 (4.0%) | 0.47( 0.14, 1.61) | 0.51 (0.14, 1.83) | 1/79 (1.3%) | 0.37 (0.05, 2.81) | 0.46 (0.05, 3.99) |
|  |  | p=0.23 | p=0.30 |  | p=0.34 | p=0.48 |
| Analysis by multilevel mixed-effects logistic regression, estimating OR (95% CI). ^a^Multivariable models adjusted for age, sex, MS phenotype, and EDSS. Abbreviations: EDSS = Expanded Disability Status Scale; MS = multiple sclerosis; OR = odds ratio; RRMS = relapsing-remitting multiple sclerosis. Results in boldface denote statistical significance (p<0.05). | | | | | | |

Table S4. Clinician-reported DMT characteristics of COVID-19 severity outcomes, confirmed COVID-19 only.

|  | Hospitalisation |  |  | ICU admission |  |  |
| --- | --- | --- | --- | --- | --- | --- |
|  | n (%) | OR (95% CI) | aOR (95% CI)^a^ | n (%) | OR (95% CI) | aOR (95% CI)^a^ |
| DMT |  |  |  |  |  |  |
| Untreated | 91/218 (41.7%) | **3.18 (1.99, 5.08)** | **1.77 (1.06, 2.94)** | 16/218 (7.3%) | 2.18 (0.86, 5.50) | 1.22 (0.46, 3.19) |
| Alemtuzumab | 3/25 (12.0%) | 0.64 (0.18, 2.34) | 0.88 (0.24, 3.22) | 1/25 (4.0%) | 1.12 (0.13, 9.84) | 1.52 (0.18, 13.13) |
| Cladribine | 2/17 (11.8%) | 0.64 (0.13, 3.09) | 0.70 (0.14, 3.52) | 0/17 (0.0%) |  |  |
| Dimethyl fumarate | 35/196 (17.9%) | 1.00 [Ref] | 1.00 [Ref] | 7/193 (3.6%) | 1.00 [Ref] | 1.00 [Ref] |
| Fingolimod | 15/138 (10.9%) | 0.63 (0.32, 1.22) | 0.64 (0.32, 1.26) | 3/137 (2.2%) | 0.62 (0.16, 2.44) | 0.68 (0.17, 2.69) |
| Glatiramer acetate | 15/70 (21.4%) | 1.27 (0.63, 2.53) | 1.02 (0.49, 2.11) | 0/70 (0.0%) |  |  |
| Interferon | 16/82 (19.5%) | 1.29 (0.65, 2.57) | 1.00 (0.49, 2.05) | 2/82 (2.4%) | 0.66 (0.13, 3.30) | 0.57 (0.11, 2.84) |
| Natalizumab | 23/160 (14.4%) | 0.82 (0.46, 1.47) | 0.80 (0.44, 1.46) | 6/159 (3.8%) | 1.02 (0.34, 3.13) | 1.06 (0.34, 3.26) |
| Ocrelizumab | 109/359 (30.4%) | **2.21 (1.42, 3.45)** | **1.61 (1.01, 2.56)** | 36/359 (10.0%) | **2.92 (1.25, 6.79)** | 2.28 (0.97, 5.35) |
| Rituximab | 61/139 (43.9%) | **4.02 (2.31, 6.99)** | **3.26 (1.84, 5.76)** | 24/139 (17.3%) | **5.68 (2.22, 14.49)** | **4.72 (1.93, 11.52)** |
| Teriflunomide | 13/72 (18.1%) | 1.21 (0.59, 2.49) | 0.89 (0.42, 1.88) | 4/72 (5.6%) | 1.60 (0.45, 5.67) | 1.26 (0.35, 4.54) |
| Other DMT | 14/45 (31.1%) | **2.28 (1.07, 4.82)** | 1.11 (0.50, 2.47) | 4/45 (8.9%) | 2.53 (0.70, 9.13) | 1.39 (0.37, 5.22) |
|  | Ventilation |  |  | Death |  |  |
|  | n (%) | OR (95% CI) | OR (95% CI)a | n (%) | OR (95% CI) | OR (95% CI)a |
| DMT |  |  |  |  |  |  |
| Untreated | 18/218 (8.3%) | 2.25 (0.89, 5.68) | 1.30 (0.48, 3.50) | 23/218 (10.6%) | **4.54 (1.67, 12.36)** | 1.48 (0.50, 4.41) |
| Alemtuzumab | 1/25 (4.0%) | 0.86 (0.10, 7.68) | 1.17 (0.13, 10.60) | 1/25 (4.0%) | 1.84 (0.20, 17.03) | 2.57 (0.23, 28.99) |
| Cladribine | 0/17 (0.0%) |  |  | 0/17 (0.0%) |  |  |
| Dimethyl fumarate | 7/193 (3.6%) | 1.00 [Ref] | 1.00 [Ref] | 5/193 (2.6%) | 1.00 [Ref] | 1.00 [Ref] |
| Fingolimod | 3/137 (2.2%) | 0.49 (0.12, 1.98) | 0.50 (0.12, 2.06) | 0/137 (0.0%) |  |  |
| Glatiramer acetate | 0/70 (0.0%) |  |  | 1/70 (1.4%) | 0.48 (0.05, 4.18) | 0.27 (0.03, 2.58) |
| Interferon | 1/82 (1.2%) | 0.23 (0.03, 1.99) | 0.19 (0.02, 1.70) | 1/82 (1.2%) | 0.59 (0.07, 5.29) | 0.33 (0.04, 3.11) |
| Natalizumab | 4/159 (2.5%) | 0.70 (0.20, 2.45) | 0.67 (0.19, 2.39) | 3/159 (1.9%) | 0.72 (0.17, 3.10) | 0.73 (0.16, 3.36) |
| Ocrelizumab | 18/359 (5.0%) | 1.37 (0.55, 3.42) | 0.99 (0.39, 2.52) | 11/359 (3.1%) | 1.11 (0.38, 3.28) | 0.47 (0.15, 1.49) |
| Rituximab | 23/139 (16.5%) | **5.43 (2.06, 14.27)** | **4.30 (1.58, 11.67)** | 5/139 (3.6%) | 2.17 (0.55, 8.55) | 1.09 (0.29, 4.16) |
| Teriflunomide | 3/72 (4.2%) | 1.05 (0.25, 4.35) | 0.66 (0.15, 2.87) | 1/72 (1.4%) | 0.58 (0.07, 5.09) | 0.32 (0.03, 3.02) |
| Other DMT | 2/45 (4.4%) | 0.92 (0.16, 5.16) | 0.47 (0.08, 2.68) | 4/45 (8.9%) | 3.95 (0.99, 15.69) | 1.17 (0.27, 5.05) |
| Analysis by multilevel mixed-effects logistic regression, estimating OR (95% CI). ^a^Multivariable models adjusted for age, sex, MS phenotype, and EDSS. Abbreviations: DMT = disease-modifying therapy; ICU = Intensive Care Unit; MS = multiple sclerosis; OR = odds ratio. Results in boldface denote statistical significance (p<0.05).  Shaded cells indicate where quantitative analyses were not possible. | | | | | | |

Table S5. Characteristics of COVID-19 severity outcomes, anti-CD20 vs pooled other DMTs, confirmed COVID-19 only.

|  | Hospitalisation |  |  | ICU admission |  |  |
| --- | --- | --- | --- | --- | --- | --- |
|  | n (%) | OR (95% CI) | aOR (95% CI)^a^ | n (%) | OR (95% CI) | aOR (95% CI)^a^ |
| Other DMT | 135/759 (17.8%) | 1.00 [Ref] | 1.00 [Ref] | 27/703 (3.8%) | 1.00 [Ref] | 1.00 [Ref] |
| Ocrelizumab | 109/336 (32.4%) | **2.21 (1.62, 3.00)** | **1.79 (1.29, 2.49)** | 36/316 (11.4%) | **3.12 (1.80, 5.41)** | **2.61 (1.52, 4.46)** |
| Rituximab | 61/131 (46.6%) | **4.02 (2.58, 6.26)** | **3.67 (2.32, 5.79)** | 24/128 (18.8%) | **6.02 (3.14, 11.55)** | **5.39 (2.94, 9.87)** |
| No DMT | 91/210 (43.3%) | **3.19 (2.27, 4.49)** | **1.97 (1.34, 2.89)** | 16/192 (8.3%) | **2.32 (1.21, 4.46)** | 1.38 (0.69, 2.75) |
| Natalizumab | 23/150 (15.3%) | 1.00 [Ref] | 1.00 [Ref] | 6/141 (4.3%) | 1.00 [Ref] | 1.00 [Ref] |
| Ocrelizumab | 109/336 (32.4%) | **2.65 (1.61, 4.37)** | **1.90 (1.13, 3.22)** | 36/316 (11.4%) | **2.89 (1.19, 7.03)** | 2.22 (0.89, 5.54) |
| Rituximab | 61/131 (46.6%) | **4.81 (2.74, 8.43)** | **4.24 (2.35, 7.64)** | 24/128 (18.8%) | **5.19 (2.05, 13.17)** | **4.29 (1.65, 11.15)** |
|  | Ventilation |  |  | Death |  |  |
|  | n (%) | OR (95% CI) | aOR (95% CI)^a^ | n (%) | OR (95% CI) | aOR (95% CI)^a^ |
| Other DMT | 21/685 (3.1%) | 1.00 [Ref] | 1.00 [Ref] | 16/745 (2.1%) | 1.00 [Ref] | 1.00 [Ref] |
| Ocrelizumab | 18/305 (5.9%) | **2.09 (1.07, 4.11)** | 1.72 (0.86, 3.46) | 11/338 (3.3%) | 1.37 (0.62, 3.03) | 0.73 (0.32, 1.70) |
| Rituximab | 23/107 (21.5%) | **8.51 (4.14, 17.48)** | **7.65 (3.63, 16.10)** | 5/124 (4.0%) | 2.66 (0.86, 8.18) | 1.72 (0.58, 5.10) |
| No DMT | 18/180 (10.0%) | **3.52 (1.79, 6.95)** | **2.35 (1.13, 4.89)** | 23/203 (11.3%) | **5.61 (2.85, 11.05)** | **2.31 (1.09, 4.90)** |
| Natalizumab | 4/136 (2.9%) | 1.00 [Ref] | 1.00 [Ref] | 3/149 (2.0%) | 1.00 [Ref] | 1.00 [Ref] |
| Ocrelizumab | 18/305 (5.9%) | 2.07 (0.69, 6.24) | 1.42 (0.45, 4.47) | 11/338 (3.3%) | 1.64 (0.45, 5.96) | 0.48 (0.11, 2.07) |
| Rituximab | 23/107 (21.5%) | **9.04 (3.02, 27.05)** | **7.40 (2.41, 22.75)** | 5/124 (4.0%) | 2.04 (0.48, 8.73) | 1.34 (0.27, 6.56) |
| Analysis by multilevel mixed-effects logistic regression, estimating OR (95% CI). ^a^Multivariable models adjusted for age, sex, MS phenotype, and EDSS. Abbreviations: DMT = disease-modifying therapy; ICU = Intensive Care Unit; MS = multiple sclerosis; OR = odds ratio. Results in boldface denote statistical significance (p<0.05). | | | | | | |

Table S6. *Severity of COVID-19 by DMT, by age, suspected+confirmed COVID-19.*

|  | Hospitalisation |  |  |  |
| --- | --- | --- | --- | --- |
|  | All persons | Age≤70 | Age>70 | Test for difference |
| DMT  Other DMT  Ocrelizumab  Rituximab  Untreated | 1.00 [Ref]  **1.75 (1.29, 2.38)**  **2.76 (1.87, 4.07)**  **2.05 (1.43, 2.94)** | 1.00 [Ref]  **1.84 (1.36, 2.50)**  **2.63 (1.78, 3.89)**  **2.27 (1.56, 3.29)** | 1.00 [Ref]  0.10 (0.01, 1.02)  -  0.76 (0.24, 2.46) | ***p=0.015***  *p=1.00*  *p=0.079* |
| Natalizumab  Ocrelizumab  Rituximab | 1.00 [Ref]  **1.86 (1.13, 3.07)**  **2.88 (1.68, 4.92)** | 1.00 [Ref]  **2.08 (1.26, 3.44)**  **2.83 (1.65, 4.87)** | 1.00 [Ref]  0.24 (0.01, 6.82)  - | *p=0.21*  *-* |
|  | ICU admission |  |  |  |
|  | All persons | Age≤70 | Age>70 | Test for difference |
| DMT  Other DMT  Ocrelizumab  Rituximab  Untreated | 1.00 [Ref]  **2.55 (1.49, 4.36)**  **4.32 (2.27, 8.23)**  1.52 (0.77, 3.02) | 1.00 [Ref]  **2.74 (1.58, 4.76)**  **4.40 (2.29, 8.48)**  1.81 (0.88, 3.70) | 1.00 [Ref]  -  -  0.48 (0.08, 2.82) | *p=0.99*  *p=1.00*  *p=0.17* |
| Natalizumab  Ocrelizumab  Rituximab | 1.00 [Ref]  2.13 (0.85, 5.35)  **3.23 (1.17, 8.91)** | 1.00 [Ref]  2.36 (0.95, 5.88)  **3.41 (1.23, 9.44)** |  |  |
|  | Artificial ventilation |  |  |  |
|  | All persons | Age≤70 | Age>70 | Test for difference |
| DMT  Other DMT  Ocrelizumab  Rituximab  Untreated | 1.00 [Ref]  1.60 (0.82, 3.14)  **6.15 (3.09, 12.27)**  **2.07 (1.01, 4.22)** | 1.00 [Ref]  1.72 (0.86, 3.42)  **6.19 (3.06, 12.52)**  **2.34 (1.10, 4.96)** |  |  |
| Natalizumab  Ocrelizumab  Rituximab | 1.00 [Ref]  1.34 (0.42, 4.24)  **5.52 (1.71, 17.84)** | 1.00 [Ref]  1.60 (0.51, 4.99)  **5.87 (1.81, 19.10)** |  |  |
|  | Death |  |  |  |
|  | All persons | Age≤70 | Age>70 | Test for difference |
| DMT  Other DMT  Ocrelizumab  Rituximab  Untreated | 1.00 [Ref]  0.76 (0.34, 1.690  1.90 (0.73, 4.93)  **2.53 (1.24, 5.15)** | 1.00 [Ref]  1.07 (0.45, 2.51)  1.82 (0.68, 4.88)  **3.89 (1.39, 16.99)** |  |  |
| Natalizumab  Ocrelizumab  Rituximab | 1.00 [Ref]  0.53 (0.13, 2.24)  1.70 (0.38, 7.62) | 1.00 [Ref]  0.92 (0.23, 3.64)  1.63 (0.38, 7.05) |  |  |
| Analysis by multilevel mixed-effects logistic regression, estimating OR (95% CI). ^a^Multivariable models adjusted for age, sex, and EDSS. Abbreviations: DMT = disease-modifying therapy; EDSS = Expanded Disability Status Scale; ICU = Intensive care unit; MS = multiple sclerosis; PR = prevalence ratio; RRMS = relapsing-remitting multiple sclerosis. Results in boldface denote statistical significance (p<0.05). Shaded cells denote where quantitative analyses not possible. | | | | |

Table S7. Severity of COVID-19 by DMT, by MS type, suspected+confirmed COVID-19.

|  | Hospitalisation |  |  |  |
| --- | --- | --- | --- | --- |
|  | All persons | RRMS | Progressive | Test for difference |
| DMT  Other DMT  Ocrelizumab  Rituximab  Untreated | 1.00 [Ref]  **1.75 (1.29, 2.38)**  **2.76 (1.87, 4.07)**  **2.05 (1.43, 2.94)** | 1.00 [Ref]  **1.96 (1.40, 2.76)**  **3.20 (2.07, 4.94)**  **1.75 (1.08, 2.82)** | 1.00 [Ref]  1.18 (0.63, 2.20)  1.66 (0.78, 3.54)  **2.16 (1.17, 3.99)** | *p=0.16*  *p=0.13*  *p=0.59* |
| Natalizumab  Ocrelizumab  Rituximab | 1.00 [Ref]  **1.86 (1.13, 3.07)**  **2.88 (1.68, 4.92)** | 1.00 [Ref]  **2.15 (1.24, 3.73)**  **3.42 (1.89, 6.18)** | 1.00 [Ref]  0.63 (0.16, 2.45)  0.88 (0.21, 3.64) | *p=0.10*  *p=0.084* |
|  | ICU admission |  |  |  |
|  | All persons | RRMS | Progressive | Test for difference |
| DMT  Other DMT  Ocrelizumab  Rituximab  Untreated | 1.00 [Ref]  **2.55 (1.49, 4.36)**  **4.32 (2.27, 8.23)**  1.52 (0.77, 3.02) | 1.00 [Ref]  **3.21 (1.68, 6.13)**  **5.84 (2.77, 12.31)**  1.71 (0.62, 4.74) | 1.00 [Ref]  1.47 (0.59, 3.68)  2.16 (0.70, 6.67)  1.05 (0.41, 2.72) | *p=0.17*  *p=0.13*  *p=0.49* |
| Natalizumab  Ocrelizumab  Rituximab | 1.00 [Ref]  2.13 (0.85, 5.35)  **3.23 (1.17, 8.91)** | 1.00 [Ref]  2.74 (0.91, 8.25)  **4.72 (1.43, 15.58)** | 1.00 [Ref]  0.75 (0.13, 4.43)  0.86 (0.12, 5.87) | *p=0.22*  *p=0.13* |
|  | Artificial ventilation |  |  |  |
|  | All persons | RRMS | Progressive | Test for difference |
| DMT  Other DMT  Ocrelizumab  Rituximab  Untreated | 1.00 [Ref]  1.60 (0.82, 3.14)  **6.15 (3.09, 12.27)**  **2.07 (1.01, 4.22)** | 1.00 [Ref]  1.52 (0.67, 3.45)  **6.57 (2.98, 14.51)**  2.24 (0.89, 5.61) | 1.00 [Ref]  1.69 (0.50, 5.69)  **5.47 (1.46, 20.52)**  1.87 (0.57, 6.17) | *p=0.89*  *p=0.81*  *p=0.82* |
| Natalizumab  Ocrelizumab  Rituximab | 1.00 [Ref]  1.34 (0.42, 4.24)  **5.52 (1.71, 17.84)** | 1.00 [Ref]  2.25 (0.48, 10.67)  **10.78 (2.28, 50.89)** | 1.00 [Ref]  0.35 (0.05, 2.22)  0.95 (0.14, 6.67) | *p=0.13*  *p=0.050* |
|  | Death |  |  |  |
|  | All persons | RRMS | Progressive | Test for difference |
| DMT  Other DMT  Ocrelizumab  Rituximab  Untreated | 1.00 [Ref]  0.76 (0.34, 1.690  1.90 (0.73, 4.93)  **2.53 (1.24, 5.15)** | 1.00 [Ref]  0.96 (0.27, 3.45)  **4.44 (1.34, 14.78)**  **3.69 (1.01, 13.51)** | 1.00 [Ref]  0.57 (0.20, 1.60)  0.62 (0.12, 3.14)  1.87 (0.80, 4.32) | *p=0.53*  *p=0.054*  *p=0.39* |
| Natalizumab  Ocrelizumab  Rituximab | 1.00 [Ref]  0.53 (0.13, 2.24)  1.70 (0.38, 7.62) | 1.00 [Ref]  0.40 (0.06, 2.66)  3.34 (0.55, 20.48) | 1.00 [Ref]  0.46 (0.04, 5.05)  0.56 (0.04, 8.40) | *p=0.94*  *p=0.28* |
| Analysis by multilevel mixed-effects logistic regression, estimating OR (95% CI). ^a^Multivariable models adjusted for age, sex, and EDSS. Abbreviations: DMT = disease-modifying therapy; EDSS = Expanded Disability Status Scale; ICU = Intensive care unit; MS = multiple sclerosis; PR = prevalence ratio; RRMS = relapsing-remitting multiple sclerosis. Results in boldface denote statistical significance (p<0.05). | | | | |

Table S8. Severity of COVID-19 by DMT, by EDSS, suspected+confirmed COVID-19.

|  | Hospitalisation |  |  |  |
| --- | --- | --- | --- | --- |
|  | All persons | EDSS 0-6 | EDSS>6 | Test for difference |
| DMT  Other DMT  Ocrelizumab  Rituximab  Untreated | 1.00 [Ref]  **1.75 (1.29, 2.38)**  **2.76 (1.87, 4.07)**  **2.05 (1.43, 2.94)** | 1.00 [Ref]  **1.76 (1.22, 2.53)**  **2.89 (1.88, 4.45)**  **2.05 (1.32, 3.19)** | 1.00 [Ref]  1.72 (0.98, 3.01)  **2.43 (1.10, 5.38)**  **2.03 (1.11, 3.71)** | *p=0.94*  *p=0.70*  *p=0.98* |
| Natalizumab  Ocrelizumab  Rituximab | 1.00 [Ref]  **1.86 (1.13, 3.07)**  **2.88 (1.68, 4.92)** | 1.00 [Ref]  1.76 (0.99, 3.12)  **2.82 (1.55, 5.11)** | 1.00 [Ref]  2.19 (0.79, 6.10)  3.09 (0.96, 9.92) | *p=0.71*  *p=0.89* |
|  | ICU admission |  |  |  |
|  | All persons | EDSS 0-6 | EDSS>6 | Test for difference |
| DMT  Other DMT  Ocrelizumab  Rituximab  Untreated | 1.00 [Ref]  **2.55 (1.49, 4.36)**  **4.32 (2.27, 8.23)**  1.52 (0.77, 3.02) | 1.00 [Ref]  **3.61 (1.81, 7.18)**  **6.01 (2.79, 12.97)**  1.99 (0.75, 5.24) | 1.00 [Ref]  1.38 (0.60, 3.16)  2.19 (0.71, 6.78)  0.98 (0.39, 2.48) | *p=0.078*  *p=0.13*  *p=0.30* |
| Natalizumab  Ocrelizumab  Rituximab | 1.00 [Ref]  2.13 (0.85, 5.35)  **3.23 (1.17, 8.91)** | 1.00 [Ref]  1.98 (0.70, 5.58)  **3.19 (1.04, 9.79)** | 1.00 [Ref]  2.71 (0.32, 22.65)  3.98 (0.40, 39.28) | *p=0.79*  *p=0.86* |
|  | Artificial ventilation |  |  |  |
|  | All persons | EDSS 0-6 | EDSS>6 | Test for difference |
| DMT  Other DMT  Ocrelizumab  Rituximab  Untreated | 1.00 [Ref]  1.60 (0.82, 3.14)  **6.15 (3.09, 12.27)**  **2.07 (1.01, 4.22)** | 1.00 [Ref]  1.38 (0.55, 3.49)  **7.08 (3.17, 15.80)**  2.22 (0.87, 5.62) | 1.00 [Ref]  1.75 (0.62, 4.94)  **4.54 (1.24, 16.56)**  1.92 (0.64, 5.79) | *p=0.74*  *p=0.56*  *p=0.84* |
| Natalizumab  Ocrelizumab  Rituximab | 1.00 [Ref]  1.34 (0.42, 4.24)  **5.52 (1.71, 17.84)** | 1.00 [Ref]  1.15 (0.28, 4.66)  **6.02 (1.57, 23.11)** | 1.00 [Ref]  1.63 (0.19, 14.06)  4.73 (0.47, 47.31) | *p=0.79*  *p=0.86* |
|  | Death |  |  |  |
|  | All persons | EDSS 0-6 | EDSS>6 | Test for difference |
| DMT  Other DMT  Ocrelizumab  Rituximab  Untreated | 1.00 [Ref]  0.76 (0.34, 1.690  1.90 (0.73, 4.93)  **2.53 (1.24, 5.15)** | 1.00 [Ref]  0.55 (0.06, 4.73)  0.93 (0.11, 8.06)  **5.05 (1.42, 17.97)** | 1.00 [Ref]  0.76 (0.31, 1.86)  2.41 (0.79, 7.42)  2.06 (0.92, 4.65) | *p=0.78*  *p=0.44*  *p=0.24* |
| Natalizumab  Ocrelizumab  Rituximab | 1.00 [Ref]  0.53 (0.13, 2.24)  1.70 (0.38, 7.62) | 1.00 [Ref]  0.19 (0.02, 2.15)  0.35 (0.03, 4.06) | 1.00 [Ref]  1.32 (0.15, 11.90)  5.49 (0.54, 55.73) | *p=0.24*  *p=0.11* |
| Analysis by multilevel mixed-effects logistic regression, estimating OR (95% CI). ^a^Multivariable models adjusted for age, sex, and MS type. Abbreviations: DMT = disease-modifying therapy; EDSS = Expanded Disability Status Scale; ICU = Intensive care unit; MS = multiple sclerosis; PR = prevalence ratio; RRMS = relapsing-remitting multiple sclerosis. Results in boldface denote statistical significance (p<0.05). | | | | |
